# Reduced Pediatric Urgent Asthma Utilization and Exacerbations During the COVID-19 Pandemic

**DOI:** 10.1101/2021.04.28.21256263

**Authors:** Jillian H. Hurst, Congwen Zhao, Nicholas S. Fitzpatrick, Benjamin A. Goldstein, Jason E. Lang

## Abstract

**Background and Objectives:** The COVID-19 pandemic has had a profound impact on healthcare access and utilization, which could have important implications for children with chronic diseases, including asthma. We sought to evaluate changes in healthcare utilization and outcomes in children with asthma during the COVID-19 pandemic.

**Methods:** We used electronic health records data to evaluate healthcare use and asthma outcomes in 3,959 children and adolescents, 5-17 years of age, with a prior diagnosis of asthma who had a history of well child visits and encounters within the healthcare system. We assessed all-cause healthcare encounters and asthma exacerbations in the 12-months preceding the start of the COVID-19 pandemic (March 1, 2019 – February 29, 2020) and the first 12-months of the pandemic (March 1, 2020 – February 28, 2021).

**Results:** All-cause healthcare encounters decreased significantly during the pandemic compared to the preceding year, including well child visits (48.1% during the pandemic vs. 66.6% in the prior year; p < 0.01), emergency department visits (9.7% vs. 21.0%; p < 0.01), and inpatient admissions (1.6% vs. 2.5%; p < 0.01), though there was over a 100-fold increase in telehealth encounters. Asthma exacerbations that required treatment with systemic steroids also decreased (127 vs. 504 exacerbations; p < 0.01). Race/ethnicity was not associated with changes in healthcare utilization or asthma outcomes.

**Conclusion:** The COVID-19 pandemic corresponded to dramatic shifts in healthcare utilization, including increased telehealth use and improved outcomes among children with asthma. Social distancing measures may have also reduced asthma trigger exposure.

## INTRODUCTION

The COVID-19 pandemic has had a profound impact on healthcare access and utilization. Many families have experienced unemployment and accompanying changes in insurance, leading to decreased or disrupted healthcare access^1^. Further, measures to prevent spread of severe acute respiratory syndrome coronavirus 2 (SARS-CoV-2), the etiological agent of COVID-19, have been implemented in a variety of settings, either broadly through stay-at-home orders and within businesses, schools, and healthcare settings, including limitations to in-person encounters, postponement of elective procedures, and viral testing prior to scheduled hospital admissions^2,3^. Patients and families have also changed the way they interact with the healthcare system, with documented increases in missed appointments and routine health maintenance such as vaccinations, and avoidance of emergency rooms and urgent care centers^4–7^. Such changes could have important implications for the health and well-being of children with chronic diseases who require regular touchpoints with the health system for disease management. Further, because comorbidities appear to increase the susceptibility to and severity of SARS-CoV-2 infections in adults, there are concerns that chronic medical conditions in children may also increase the risk of COVID-19^8^.

Asthma is the most common chronic condition among children less than 18 years of age, with more than 5.5 million children and adolescents in the United States estimated to have an asthma diagnosis^9^. Childhood asthma is primarily managed through regular assessment and monitoring, control of factors that aggravate symptoms, pharmacologic therapy, and education in symptom management and treatment^10^. Notably, all components require regular interactions with the health system to ensure timely evaluation and provision of prescriptions for controller and rescue medications. We and others have previously reported that children who have yearly well child visits have decreased risk of asthma exacerbations and related emergency department visits compared with children who do not have regular well child visits^11^. Similarly, continuity of care is associated with improved asthma outcomes in children and adolescents^12–16^. Taken together, these findings suggest that disruptions to healthcare access and utilization patterns may result in increased exacerbations and poor outcomes in children with asthma.

Because the COVID-19 pandemic has significantly altered healthcare provision and access, we hypothesized that the pandemic would markedly alter care patterns and health outcomes in children and adolescents with asthma. Further, stay-at-home orders, school closures, and other social distancing measures could alter exposures to common asthma triggers, including allergens and respiratory viruses. We previously characterized a cohort of over 5000 children and adolescents with asthma who receive primary care through an academic medical center-based healthcare system. Herein, we conducted pre-post cohort study using electronic health records (EHR) data to evaluate healthcare utilization and asthma outcomes in children and adolescents in the 12 months prior to the beginning of the COVID-19 pandemic (March 1, 2019 – February 29, 2020) and the first year of the pandemic (March 1, 2020 – February 28, 2021).

## METHODS

### Study design

This study was evaluated by the Duke University Health System (DUHS) Institutional Review Board (Pro00091342). The requirement for informed consent was waived under 45 CFR 46.116. The DUHS is a comprehensive medical system that includes a large tertiary care hospital, which includes a children’s health center and hospital, two community hospitals, and a network of primary and urgent care pediatric and family medicine clinics and inpatient and outpatient specialty services. DUHS is the primary care provider within Durham County, North Carolina, and is estimated to provide healthcare services, including primary and specialty care, for over 85% of Durham residents. We abstracted patient records from the DUHS EHR system to conduct a retrospective cohort study to assess changes in healthcare use and asthma outcomes in children and adolescents with asthma who receive primary care through the DUHS.

### Participant selection

Patients included in the cohort were required to meet the following criteria: 1) ages 5 years to <18 years of age as of March 1, 2020; 2) previous asthma diagnosis; 3) home address within Durham County, North Carolina for the study period (March 1, 2019 through February 28, 2021); 4) at least one in-person healthcare encounter of any type in the year prior to the start of the COVID-19 pandemic (January 1, 2019 through December 31, 2019) (**Figure 1**). Patients were excluded from the study cohort if they were under or over the specified age range during the study period or had a recorded residential address outside of Durham County during the study period. We previously reported the development and validation of a computable phenotype to identify children with asthma diagnoses, which we used to select patients for this study^17^. To meet the criteria for study-defined asthma, patients had to have (1) either an asthma-appropriate International Classification of Diseases, 10^th^ Revision, (ICD-10) (J45.x) diagnosis code documented in the EHR problem list section or 2 or more outpatient or ED encounters (or 1 hospital encounter) with the appropriate ICD-10 coding and (2) documented prescription of 1 or more asthma medications.

**Figure 1.**
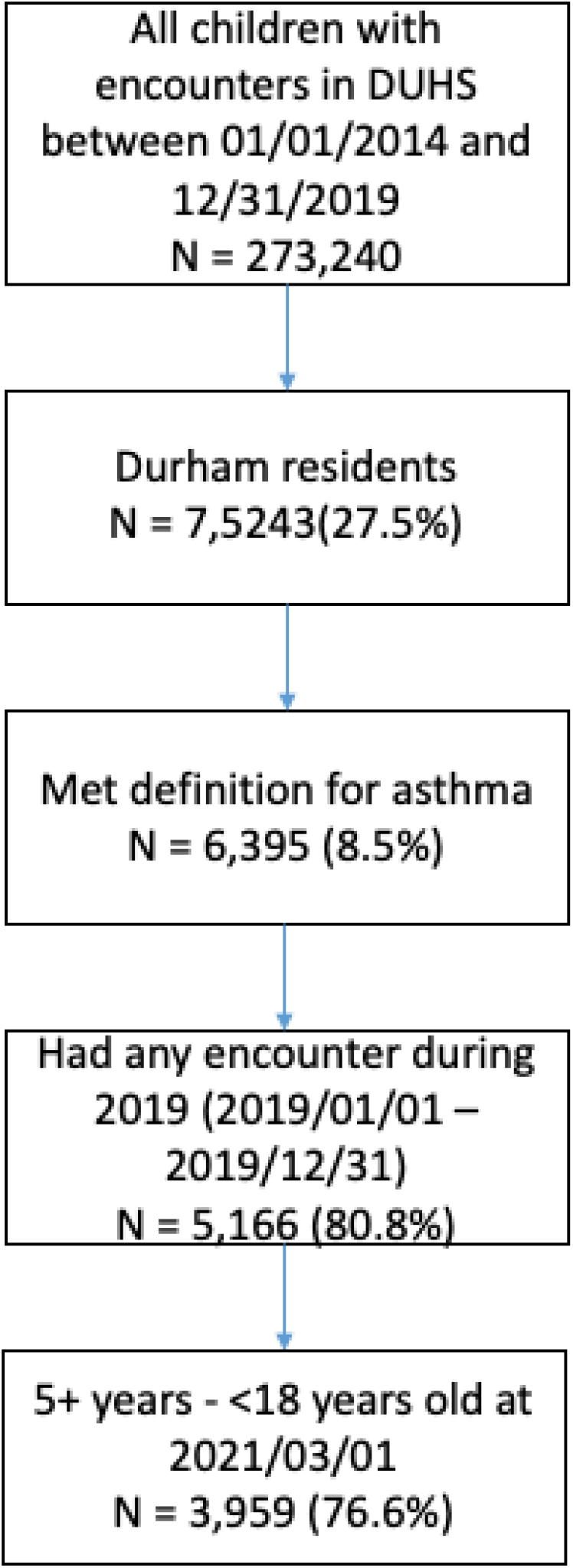
Consolidated Standards of Reporting Trials Diagram.

We abstracted information on patient age, race and ethnicity (categorized as non-Hispanic white, non-Hispanic black, Hispanic and other), primary payor (based on most recent encounter to the March 1, 2020 – the study midpoint), number and types of healthcare encounters within the DUHS, asthma exacerbations that required medical attention, SARS-CoV-2 testing and results, and antibiotic prescriptions.

### Outcomes of interest

We evaluated changes in healthcare utilization and asthma outcomes in the year prior to the COVID-19 pandemic (“Year 1”: March 1, 2019 – February 29, 2020) and the first year of the COVID-19 pandemic (“Year 2”: March 1, 2020 – February 28, 2021). The first outcomes of interest were the number and types of all-cause healthcare visits before and during the COVID-19 pandemic, including: well child visits, defined as scheduled primary care office-based encounters or initial consults that also had a diagnosis code representing routine health supervision (z00.110’, ‘z00.111’, ‘z00.121’, or ‘z00.129’); hospital encounters greater than 24 hours in duration; hospital or emergency department encounters less than 24 hours in duration; and all other outpatient encounters, including urgent care visits, telehealth encounters, and other encounters that do not meet criteria for the three prior categories. Additionally, we evaluated asthma-related healthcare utilization and the occurrence of asthma exacerbations, which were defined as encounter type with an asthma diagnosis code (J45.x) and the administration or prescription for an oral corticosteroid. Because upper respiratory infections, including SARS-CoV-2, could potentially contribute to asthma exacerbations, we assessed antibiotic prescriptions, the number of SARS-CoV-2 PCR tests, and the number of positive SARS-CoV-2 tests within our population. We also assessed potential impacts of stay-at-home and social distancing measures with respect to asthma exacerbation rates during the first year of the COVID-19 pandemic; state-wide social distancing and stay-at-home orders described in **Supplemental Table 1**. Lastly, in order to evaluate potential access disparities during the COVID-19 pandemic, we assessed healthcare encounters by race/ethnicity for encounters before and after the start of the pandemic.

### Statistical analyses

The primary objective of the study was to assess the impact of the COVID-19 pandemic on healthcare utilization and asthma-related healthcare encounters and outcomes among children with asthma. We first described baseline patient characteristics. For our primary analysis, we assessed the type and number of healthcare encounters during the pre-pandemic year and the first year of the COVID-19 pandemic, reporting the number of encounters, percent of patients with a particular encounter type reported, and the median and interquartile range (IQR) for the number of visits for a given encounter type. We used a paired Wilcoxon test to assess differences in healthcare utilization in the pre-pandemic year compared to the first year of the COVID-19 pandemic. The same approaches were used to evaluate differences in asthma-associated healthcare encounters and asthma exacerbations. Data management and analyses were performed with R version 3.6.1.

## RESULTS

### Patient Characteristics

We previously identified 5,656 children 5 to <18 years of age who met the criteria for asthma diagnosis, had a Durham County address of residence, and were identified as receiving primary care through the DUHS^11^. Of this initial cohort, 3,959 had at least one in-person healthcare encounter in the year prior to the start of the COVID-19 pandemic (Year 1) and were therefore included in the primary analysis. The demographic characteristics of this cohort are shown in **Table 1**. The median age of the population as of March 1, 2020, was 11 years of age; there was a slightly greater number of males than females, in line with the reported higher prevalence of asthma in male compared with female children and adolescents^18^. Nearly 60% of children and adolescents in the cohort were publicly insured and over half of the patients identified as non-Hispanic Black or African American.

**Table 1.** Baseline characteristics of the eligible cohort (N=3,959)

| **Table 1.** Baseline characteristics of the eligible cohort (N=3,959) |  |  |
| --- | --- | --- |
| **Characteristics** | **N or Median** | **IQR or %** |
| **Age** | 11 | (9, 14) |
| **Sex** | **N** | **%** |
| Female | 1,667 | 42.1% |
| Male | 2,292 | 57.9% |
| **Race/Ethnicity** | **N** | **%** |
| Hispanic | 574 | 14.5% |
| Non-Hispanic Black | 2,209 | 55.8% |
| Non-Hispanic White | 794 | 20.1% |
| Other/Unknown | 382 | 9.6% |
| **Insurance type** | **N** | **%** |
| Private | 1,487 | 37.6% |
| Public | 2,343 | 59.2% |
| Other/Unknown | 129 | 3.2% |
| Tested for SARS-CoV-2 | 741 | 18.7% |
| Tested positive for SARS-CoV-2 infection | 125 | 3.2% |

### Overall healthcare utilization in the year before versus during the COVID-19 pandemic

We first evaluated overall healthcare utilization in the year before and the first year of the COVID-19 pandemic, including preventive healthcare (well child visits) and visits for acute care (**Table 2**). In Year 1, patients in the cohort had a median of 6 healthcare encounters, and 66.6% had a well child visit. During Year 2, the median visits declined to 5 per patient and 48.1% had a well child visit (p < 0.01). Notably, the percentages of patients who had inpatient admissions, emergency department visits, and other types of outpatient encounters all declined significantly during Year 2 compared to the Year 1 (p <0.01); however, telehealth visits increased significantly. In Year 1, 11 patients (0.03%) had a telehealth visit, while in Year 2, 1,385 (35%) patients had telehealth visits.

**Table 2.** All causes healthcare utilization in the year before and the year during the COVID-19 pandemic

|  |  |  |  |  |  |  |  |  |  |
| --- | --- | --- | --- | --- | --- | --- | --- | --- | --- |
| Encounter types | Pre-pandemic (03/01/2019 – 02/29/2020) |  |  |  | During pandemic (03/01/2020 – 02/28/2021) |  |  |  | *P* value |
|  | N | Percentage | Median | IQR | N | Percentage | Median | IQR |  |
| All healthcare encounters | 3,882 | 98.1% | 6 | (3, 10) | 3,307 | 83.5% | 5 | (3, 8) | <0.01 |
| Well child visits | 2,635 | 66.6% | 1 | (1, 1) | 1,905 | 48.1% | 1 | (1, 1) | <0.01 |
| Telehealth visits | 11 | 0.3% | 1 | (1, 1.5) | 1,385 | 35.0% | 1 | (1, 2) | <0.01 |
| Outpatient visits (excluding telehealth) | 3,818 | 96.4% | 5 | (3, 9) | 3,187 | 80.5% | 4 | (2, 6) | <0.01 |
| Emergency department | 833 | 21.0% | 1 | (1, 2) | 384 | 9.7% | 1 | (1, 2) | <0.01 |
| Inpatient admissions | 97 | 2.5% | 1 | (1, 1) | 62 | 1.6% | 1 | (1, 1) | <0.01 |

### Asthma exacerbations and asthma-related healthcare use before and during the COVID-19 pandemic

We next evaluated changes in asthma outcomes and healthcare utilization related to asthma exacerbations. In Year 1, 504 patients in the cohort (12.7%) had an asthma exacerbation. We observed a significant decrease in the number of asthma exacerbations among patients in this cohort in Year 2, with only 127 patients (3.2%) having an exacerbation recorded. All healthcare encounters with asthma exacerbations decreased (**Table 3**).

**Table 3.** Asthma-related healthcare utilization in the year before and the year during the COVID-19 pandemic

|  |  |  |  |  |  |  |  |  |  |
| --- | --- | --- | --- | --- | --- | --- | --- | --- | --- |
| Encounter types | Pre-pandemic (03/01/2019 – 02/29/2020) |  |  |  | During pandemic (03/01/2020 – 02/28/2021) |  |  |  | *P* value |
|  | N | Percentage | Median | IQR | N | Percentage | Median | IQR |  |
| All asthma exacerbations | 504 | 12.7% | 1 | (1, 2) | 127 | 3.2% | 1 | (1, 1) | <0.01 |
| Outpatient encounters | 399 | 10.1% | 1 | (1, 1) | 89 | 2.2% | 1 | (1, 1) | <0.01 |
| ED or hospital encounters < 24 hrs | 112 | 2.8% | 1 | (1, 1) | 34 | 0.9% | 1 | (1, 1) | <0.01 |
| Hospital encounters > 24 hrs | 42 | 1.1% | 1 | (1, 1) | 11 | 0.3% | 1 | (1, 1) | <0.01 |
| Antibiotic prescriptions | 283 | 7.1% | 1 | (1, 1) | 106 | 2.7% | 1 | (1, 1) | <0.01 |
| SARS-CoV-2 tests administered | --- | --- | --- | --- | 741 | 18.7% | --- | --- | --- |
| Positive SARS-CoV-2 tests | --- | --- | --- | --- | 125 | 3.2% | --- | --- | --- |

Asthma exacerbations in children exhibit seasonal peaks that coincide with fall return to school and winter respiratory infections ^19–21^. In order to assess whether these seasonal patterns continued during the first year of the pandemic (Year 2), we plotted the number of asthma exacerbations by month for each year (**Figure 2**). In the pre-pandemic year (Year 1), the month of July had the lowest number of exacerbations, followed by June and August, while the greatest number of exacerbations were observed in May, December, and April. During the pandemic year, exacerbations during all months were significantly lower, with the greatest number of exacerbations occurring in March, at the very start of the pandemic, while the number of exacerbations was markedly lower the rest of the year. Decreases in asthma exacerbations coincided with initial lockdown measures in March, with slight increases in exacerbations appearing to coincide with reopening measures in May and September (**Figure 2 and Supplemental Table 1**).

**Figure 2.**
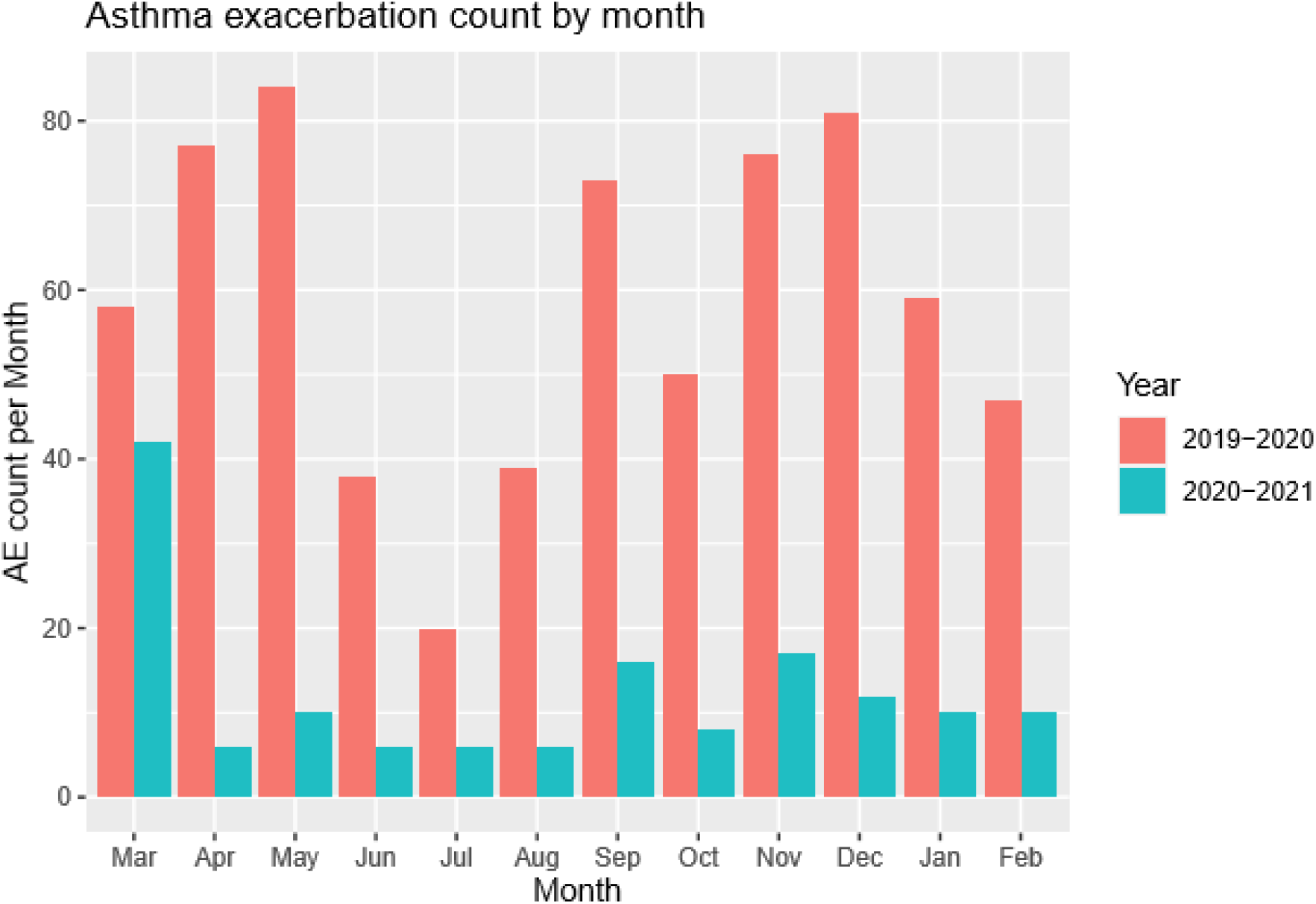
Number of asthma exacerbations per month in the year prior to and the first year of the COVID-19 pandemic. The number of asthma exacerbations (AEs) associated with a healthcare encounter in which systemic steroids were prescribed or administered was plotted by month. Data for the year prior to the COVID-19 pandemic (March 1, 2019 – February 29, 2020) are shown in red; data for the first year of the COVID-19 pandemic (March 1, 2020 – February 28, 2021) are shown in blue.

We also evaluated markers of respiratory infections within the cohort, including antibiotic prescriptions and SARS-CoV-2 tests. There was a significant decline in the number of antibiotic prescriptions in Year 2 compared to Year 1 (283 in year 1 compared to 106 in year 2, p<0.01), suggesting a potential decrease in infections that could contribute to asthma exacerbations. A total of 741 (18.7%) of the 3959 children within the asthma cohort were tested for SARS-CoV-2. Among those tested, 125 (16.9%) were positive for SARS-CoV-2. None of the children with asthma within this cohort were hospitalized due to COVID-19.

### Effect of race/ethnicity on healthcare utilization and asthma exacerbations

We did not find any significant differences in healthcare use or in the number of asthma exacerbations based on race/ethnicity (**Supplemental Figure 1**).

## DISCUSSION

We evaluated changes in healthcare utilization for children and adolescents with asthma before and after the start of the COVID-19 pandemic. We found that overall healthcare utilization decreased in all settings, except for telehealth encounters, which increased dramatically during the COVID-19 pandemic. Importantly, asthma outcomes for the children and adolescents in this cohort were improved during the first year of the pandemic, with significant reductions in asthma exacerbations and urgent asthma-related healthcare encounters. Importantly, these asthma improvements were apparent in children of all races/ethnicities, suggesting that there were not disparities in access to care related to the COVID-19 pandemic during the study period.

The COVID-19 pandemic has had a significant impact on healthcare utilization patterns among children and adolescents, generating concerns related to health outcomes for children with chronic diseases, including asthma. A retrospective analysis of data from 18,912 pediatric patients with asthma treated in a large healthcare network in California found reductions of 68-90% in asthma-related hospitalizations and emergency department visits during the first six months of the pandemic compared to historical rates from 2017-2019^22^. Notably, improvements in asthma-related emergency department visits during the pandemic did not extend to Black children; however, there prescriptions for oral corticosteroid prescriptions decreased in this population during the pandemic^22^. Additionally, the study found a greater increase in telehealth visits among publicly insured children compared to those with private insurance. An analysis of data from 77 pediatric intensive care unit (PICU) sites in the United States during the first six months of the pandemic found a marked decrease in admissions related to asthma, with 1,327 patients admitted during Quarter 2 of 2019 (pre-pandemic) compared to 241 patients in Quarter 2 of 2020 (during the pandemic)^23^. Another retrospective analysis using data from 1054 children with asthma and 505 non-asthmatic children in the multinational Pediatric Asthma in Real Life (PeARL) cohort identified decreased asthma-related hospitalizations and emergency department visits and improved scores on validated asthma control measures, though there was an increase in the frequency of treatment escalation^24^. Similar to the studies described above, we also identified a marked decrease in hospitalizations, emergency department visits, and asthma exacerbations during the first year of the pandemic compared to the prior year.

The reasons underlying improved asthma control and decreased acute care encounters for pediatric asthma are likely multifactorial. The PeARL study reported decreased incidence of upper and lower respiratory infections and febrile episodes among children with and without asthma, suggesting that decreased infectious triggers could play a role in improved asthma outcomes^24^. Supporting the potential role of decreased respiratory infections, a single center study of children with asthma reported the absence of the typical fall seasonal spike in asthma exacerbations that has been attributed to increased circulation of common respiratory viruses associated with schools reopening, as well as increased exposure to indoor allergens, changes in outdoor allergens, and colder weather^25^. While we did not have access to data on incidence of respiratory infections, we did find a decrease in prescriptions for antibiotics commonly prescribed for respiratory infections, suggesting decreased infections in our study population.

Moreover, there have been multiple epidemiological studies in multiple countries demonstrating decreases in rates of rhinoviruses, influenza, and respiratory syncytial virus that have been attributed to masking and social distancing measures^26,27^. We also observed slight increases in asthma exacerbation rates that coincided with the timing of reopening orders, suggesting that reduced social distancing could have potentially been a contributing factor.

Our data demonstrate that telehealth visits increased significantly for children with asthma during the COVID-19 pandemic. Though telehealth visits was available prior to the pandemic, only 15% of pediatricians reported using this visit modality prior to the pandemic^28^. An analysis of EHR data from 45 pediatric primary care practices showed rapid uptake of telehealth within the first month of initiation of stay-at-home orders, and continued use through May 2020, with the majority of visits pertaining to mental health and dermatological concerns^29^. Telehealth visits were inversely correlated with the number of in-person office visits, and the number of total encounters remained closer to pre-pandemic levels in practices using telehealth. A more recent study demonstrated associations between telehealth visit use and stay-at-home orders and phased reopening stages in pediatric practices^30^. In addition to telehealth, remote lung function evaluation, including home spirometry, has become more common during the COVID-19 pandemic; however, due to device and cost availability, this practice is not yet widespread^31,32^.

## STRENGTHS AND LIMITATIONS

Our study had several strengths. Our cohort included nearly 4000 children and adolescents who were established patients with the DUHS prior to the start of the COVID-19 pandemic, allowing assessment of changes in healthcare use and outcomes within a well-characterized population.

We performed our analyses in a setting in which the patients receive the majority of their care within a single system, increasing confidence that all healthcare interactions have been captured for the population. Additionally, the population under study is highly diverse, providing an opportunity to evaluate potential inequities in healthcare and asthma outcomes. Despite these strengths, our study had several limitations. This is a single center study, so all findings may not be generalizable. We used an asthma exacerbation definition that required the prescription or administration of systemic steroids and therefore did not capture mild exacerbations. Further, we did not have access to asthma control or lung function data and cannot make any conclusions about overall asthma control. Finally, we used the prescription of antibiotics commonly used for respiratory infections as a surrogate for respiratory infection rates within the population, but do not have a more direct measure of respiratory infections or other potential exposures that could contribute to asthma outcomes within this population.

## CONCLUSIONS

We conducted an analysis of healthcare utilization and asthma outcomes in children before and during the COVID-19 pandemic. We found that all encounter types decreased during the first year of the pandemic, with a compensatory increase in telehealth visits. Importantly, we observed a significant decrease in asthma exacerbations and associated healthcare encounters, suggesting that conditions during the pandemic contributed to improved asthma outcomes in children. Moreover, improvements were seen across the full population, regardless of race/ethnicity, indicating that there were not disparities in telehealth healthcare access or asthma outcomes. Future work should seek to fully evaluate the specific factors that contributed to improved outcomes in children with asthma, particularly with respect to factors that could be leveraged after the end of the COVID-19 pandemic, including the use of telehealth for the provision of care.

## Data Availability

Deidentified analytic sets will be provided upon request to the authors.

## Notes

**Funding/Support:** Funded by a grant from the National Heart, Lung, and Blood Institute (5R21HL145415-02, JEL/BAG) and by the Duke Children’s Health & Discovery Initiative. The project was supported by the National Center for Advancing Translational Sciences (NCATS), National Institutes of Health (NIH), through Grant Award Number UL1TR001117 at Duke University.

### Competing Interest Statement

The authors have declared no competing interest.

### Clinical Trial

This was a retrospective data analysis.

### Funding Statement

Funded by a grant from the National Heart, Lung, and Blood Institute (5R21HL145415-02, JEL/BAG) and by the Duke Children’s Health & Discovery Initiative. The project was supported by the National Center for Advancing Translational Sciences (NCATS), National Institutes of Health (NIH), through Grant Award Number UL1TR001117 at Duke University.

### Author Declarations

This study was evaluated by the Duke University Health System (DUHS) Institutional Review Board (Pro00091342).

