## Supplemental Table 1 for "Reduced Pediatric Urgent Asthma Utilization and Exacerbations During the COVID-19 Pandemic"

**SUPPLEMENTAL DATA**

| **Supplemental Table 1.** North Carolina Social Distancing and Shelter-in-Place Orders During the Study Period* | |
| --- | --- |
| **Date of State Order** | **Order Type** |
| March 10, 2020 | Declaration of a state of emergency |
| March 14, 2020 | K-12 schools closed state-wide |
| March 17, 2020 | Restaurants and bars closed for dine-in service |
| March 23, 2020 | Mass gatherings of greater than 50 individuals banned |
| March 27, 2020 | State-wide Stay-at-Home order issued (Phase 1) |
| May 20, 2020 | Initial Stay-at-Home order is replaced with a relaxed Safer-at-Home order (Phase 2) |
| June 24, 2020 | State-wide requirement for face coverings in public indoor spaces if a distance of at least 6-feet of distance between individuals cannot be maintained |
| September 4, 2020 | State moves to Phase 2.5 of reopening |
| September 30, 2020 | State moves to Phase 3 of reopening |
| November 23, 2020 | State-wide requirement for face coverings in both indoor settings regardless of social distancing and in outdoor settings if at least 6-feet of distance between individuals cannot be maintained |
| December 8, 2020 | State-wide modified Stay-at-Home order, including a night-time public closure period for certain businesses |
| February 24, 2021 | State-wide modified Stay-at-Home order lifted; state returns to Phase 3 |

**Detailed information regarding orders is available at* [*https://www.nc.gov/covid-19/covid-19-orders-directives*](https://www.nc.gov/covid-19/covid-19-orders-directives)

**Supplemental Figures and Figure Legends**

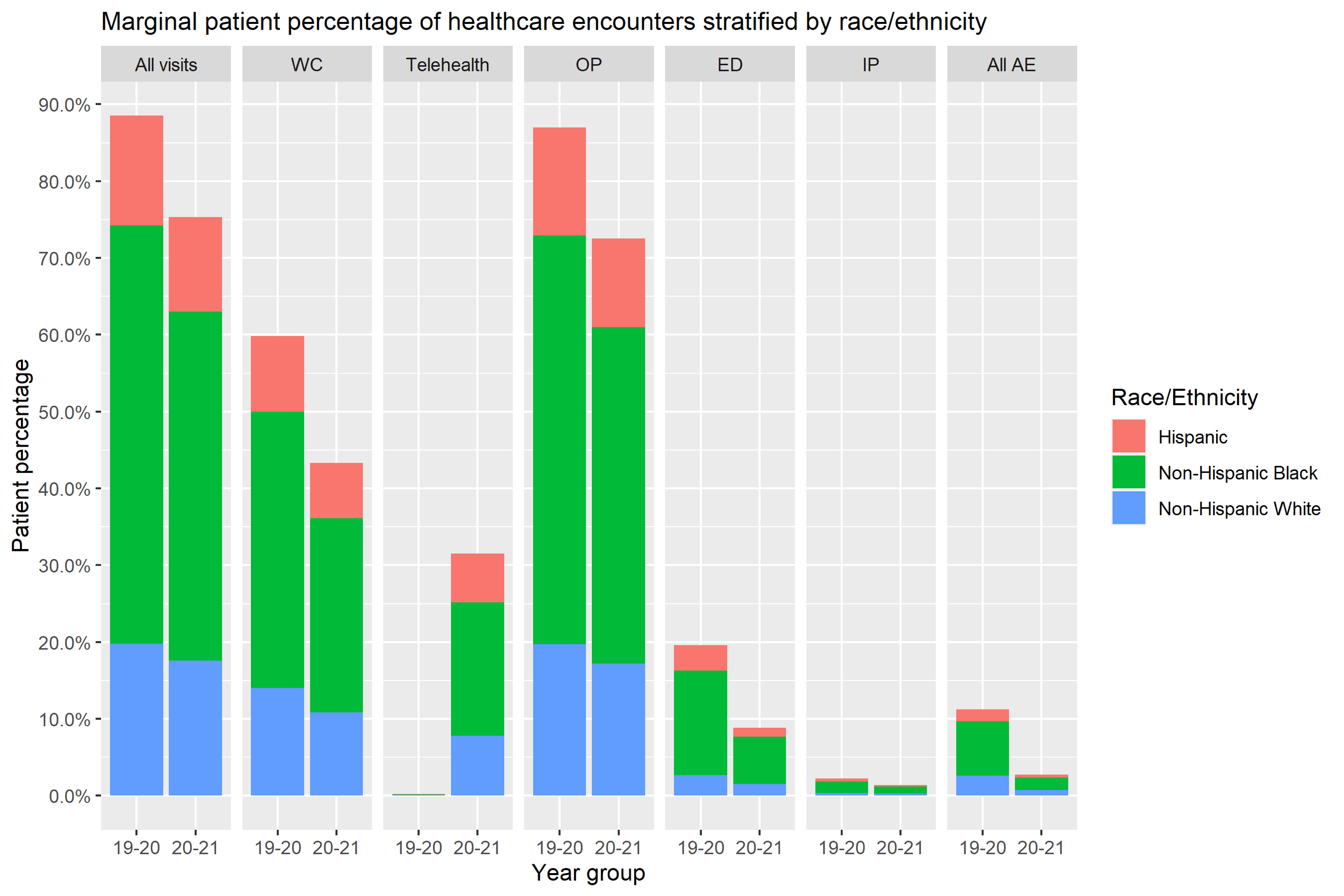
**Supplemental Figure 1.** Number and type of healthcare encounters and asthma exacerbations in the year prior to and the first year of the COVID-19 pandemic, stratified by race/ethnicity.

The percentage of patients who identified as non-Hispanic white, non-Hispanic Black/African American, or other races/ethnicities were plotted for each type of healthcare encounter, including all encounters, well child visits (WC), telehealth visits (Telehealth), other types of outpatient visits (OP), emergency department visits (ED), inpatient admissions (IP), and all asthma exacerbations (AE). Data are shown for the year prior to the COVID-19 pandemic (19-20) and the first year of the COVID-19 pandemic (20-21).
